# Characterizing the spatiotemporal heterogeneity of the COVID-19 vaccination landscape

**DOI:** 10.1101/2021.10.04.21263345

**Authors:** Andrew Tiu, Zachary Susswein, Alexes Merritt, Shweta Bansal

## Abstract

It is critical that we maximize vaccination coverage across the United States so that SARS-CoV-2 transmission can be suppressed, and we can sustain the recent reopening of the nation. Maximizing vaccination requires that we track vaccination patterns to measure the progress of the vaccination campaign and target locations that may be undervaccinated. To improve efforts to track and characterize COVID-19 vaccination progress in the United States, we integrate CDC and state-provided vaccination data, identifying and rectifying discrepancies between these data sources. We find that COVID-19 vaccination coverage in the US exhibits significant spatial heterogeneity at the county level and statistically identify spatial clusters of undervaccination, all with foci in the southern US. Vaccination progress at the county level is also variable; many counties stalled in vaccination into June 2021 and few recovered by July, with transmission of the Delta variant rapidly rising. Using a comparison with a mechanistic growth model fitted to our integrated data, we classify vaccination dynamics across time at the county scale. Our findings underline the importance of curating accurate, fine-scale vaccination data and the continued need for widespread vaccination in the US, especially in the wake of the highly transmissible Delta variant.

## Introduction

The rapid development of multiple effective vaccines for COVID-19 has been an essential pharmaceutical response to the COVID-19 pandemic, reducing transmission and severe disease. However, vaccine development is but a single step in achieving COVID-19 suppression which requires widespread vaccination. As of August 1, 2021, on average, one in every two Americans has full vaccine protection against SARS-CoV-2. However, a highly decentralized approach to mass vaccination led by individual states has created a patchwork of vaccine uptake, leading to a highly heterogeneous landscape of vaccine immunity [1, 2, 3]. Alongside the relaxation of social distancing restrictions and other non-pharmaceutical interventions, the rising prevalence of the highly transmissible Delta (B.1.617.2) variant has significantly raised the threat posed by COVID-19, causing a surge in cases and mortality especially in areas with low vaccination rates [4, 5]. Additionally, while vaccination offers strong protection against serious illness, breakthrough infections associated with the Delta variant can occur in vaccinated individuals [6, 7], placing even greater urgency on increasing vaccination levels in the US [8, 9]. These factors emphasize the need to closely monitor US vaccination progress and characterize variation in vaccination rates across both geography and time.

A variety of COVID-19 vaccination trackers exist, presenting both state and county-level information [10, 11, 12] based on data from the Centers for Disease Control and Prevention (CDC), which in turn relies on data reported by state and local health departments. Granular vaccination data were not available from the CDC until four months into the US COVID-19 vaccination campaign, and significant data missingness and incompleteness persist in these data a year into the campaign. These issues lead to misleading estimates of vaccination distribution and impede efforts to measure vaccination progress and identify target locations that may be undervaccinated.

In addition to tracking the vaccination campaign, a better understanding of the spatiotemporal distribution of historical COVID-19 vaccination coverage is critical to quantify population immunity [13] and estimate future transmission potential [1]. Spatial heterogeneity in vaccination can enable outbreaks, as clusters of unvaccinated individuals can cause resurgence despite high overall vaccination rates (e.g., in the case of measles [14, 15, 16, 17] and pertussis [18, 19]), despite high vaccination coverage at the national level. Given the importance of spatial heterogeneity, tracking vaccination at a fine spatial scale is also critical as vaccine uptake can vary widely within and between large geographical areas (e.g., in the case of influenza [20]) and large-scale aggregation of vaccination metrics can mask local vulnerabilities [14, 21, 22, 23].

A spatiotemporal characterization of US COVID-19 vaccination remains limited. Previous work has investigated temporal trends up to May 2021 in US vaccination rates among adults, finding that vaccination lagged in younger age groups even when timing of vaccine eligibility was accounted for [24]. Studies have also found evidence for county-level spatial disparities in vaccination and have linked them to social vulnerability, with some studies showing counties of lower socioeconomic status having lower vaccination coverage [25, 26], while others show higher vaccination rates in counties with high educational attainment and high proportion of minority residents [27]. While these studies provide a large-scale, early analysis of COVID-19 vaccination patterns in the US, they suffer from data missingness, analyze partial vaccination patterns in some cases, or they do not capture the entire trajectory of the vaccination campaign, particularly in light of Delta transmission.

Here, we characterize the US COVID-19 vaccination landscape at the county scale over time. We integrate state and local vaccination data with CDC-provided data to improve data coverage and accuracy. We find spatial clusters of low vaccination counties and examine these clusters across time. Additionally, we characterize the temporal dynamics of vaccination at the county scale, and compare the observed dynamics to a null model to describe the processes underlying vaccination progress. Our findings retrospectively provide an understanding of the arc of vaccine uptake to guide decision-making on sustaining vaccine confidence, and prospectively inform timely decisions about outbreak risk and variant emergence.

## Methods

### Data collation and cleaning

To characterize US county-level COVID-19 vaccination patterns accurately we integrate data from the CDC with data provided by state health departments. We collect data on complete vaccination– the number of county residents that were vaccinated fully (with one dose of the Jannsen vaccine or two doses of the Moderna or Pfizer vaccine).

The CDC reports vaccination data in the COVID Data Tracker Integrated County View [28, 29]. County-level complete vaccination data are available for all 50 states except Texas and Hawaii; additionally, there are counties with no data available in California, Virginia, and Massachusetts. The state vaccination data come from each state’s health department and have been downloaded in a machine-readable format, scraped from the health department website, or scraped from data aggregators [30]. We also supplement with data collection from other academic researchers [31] and verify against available state data. (See Supplementary Table S1 for details.)

We compare the CDC-reported vaccination counts to those provided by state health departments to identify discrepancies. For states where the CDC-reported counts are smaller than those reported by the state, we use the state-reported data for the corresponding dates (Supplementary Table S1) as advised by the CDC. We also identify counties where the CDC-reported vaccination counts are higher than those reported by the state. Most of these discrepancies can be explained by the presence of federally-serviced populations for which the CDC reports data but the state does not. In these cases, we use the CDC-reported data for the county in question (Supplementary Table S2). For more details, see the Supplement.

We collate these disparate data sources to produce a single estimate of cumulative vaccination counts for every county. We use population estimates from the 2019 American Community Survey of the US Census to produce vaccination coverage estimates as cumulative vaccination counts divided by total population size for each county. This is important as it makes our vaccination coverage estimates (a) comparable across the entire country (in contrast to coverage measures based on a variety of population size estimates used by each state); and (b) epidemiologically-relevant as we capture the entire susceptible population size (in contrast to coverage measures reported as a proportion of eligible population sizes of different ages).

### Spatial heterogeneity & clustering

To characterize the spatial structure in vaccination patterns, we analyze our collated vaccination coverage estimates with standard spatial statistic techniques. First, we calculate Moran’s I to characterize the spatial autocorrelation in county-level vaccination coverage for each week of 2021.

Second, we use Kulldorff’s Poisson spatial scan statistic (SaTScan v10.0) to detect clusters of low vaccination US counties [32, 33]. Complete vaccination counts for the week ending August 1, 2021 in each county are assumed to be Poisson distributed, with county locations defined by their centroids. For each county, expected vaccination counts are calculated as the product of the overall vaccination rate and county population, forming the null hypothesis. For each county, a circular spatial window (scanning window) centered at that location is constructed and the expected vaccination case counts are compared to the observed counts. If there is a lower number of cases than expected within the window, the likelihood ratio is calculated (or defined as 1 otherwise). The radius of the scanning window is then incrementally increased, to include neighboring locations, up to a user defined limit (which we set to 1% of the US population size). This process is repeated for all considered counties and window sizes. The reported clusters are chosen to have no geographical overlap with each other. For each cluster, p-values are calculated via Monte Carlo hypothesis testing, generating replicates of the data under the null hypothesis of uniform probability of vaccination across counties.

To examine how low vaccination clusters persist or change over time, we repeat the above spatial analysis for the first complete week of each month from January 2021 to August 2021, for a total of eight weeks.

### Temporal dynamics

To identify periods of stalling in vaccination rates before the predominance of the Delta variant, we use a threshold of 0.4% on the weekly growth rate of observed vaccination coverage (amounting to 40 new vaccinated individuals per week in rural counties ∼ 10, 000 population). We focus on the period from the week of May 30, 2021 (week 21) to the week of June 27, 2021 (week 25).

To identify vaccination growth following widespread outbreaks of the Delta variant, we estimate the average growth rate during the period of week 26 to week 30. Growth is considered significant if it is larger than 0.4% of the population size.

### Model-based comparison

We use a simple growth model as a reference point against which to compare county vaccination patterns across the country. We fit partially pooled county-level logistic growth models to observed vaccination rates from the week of January 10, 2021 (week 1) to the week of June 27, 2021 (week 25), for a total of 25 weeks. The logistic growth model is a classic ecological model representing initial exponential population growth, a gradual decrease in the growth rate, and, in the limit, the approach to an asymptotic maximum population size. The partial pooling structure allows for systematic county-level deviation from overall state trends, while still sharing information across counties. We fit these nonlinear mixed-effects models with a Bayesian approach. Each county-specific parameterization is drawn from a hierarchical state-level distribution, such that information is shared across counties within states.

In county *i* within state *S* at time *t*, we estimate complete vaccination rate *y*_*it*_, asymptotic vaccination rate *α*_*i*_, intrinsic growth rate of vaccination *β*_*i*_, and inflection time point *γ*_*i*_. (For more details, see the Supplement).

To determine if a weekly observation deviates from our model during the second stage of the vaccination campaign (defined as beyond week 15), we identify if the observation is part of the latest and longest period of consecutive observations of at least two weeks that falls outside of the 95% prediction interval.

## Results

To track US COVID-19 vaccination progress at a fine scale over time, we integrate county-level vaccination data from the CDC and state health departments, allowing us to account for and correct discrepancies between data sources. Using these data, we characterize the spatiotemporal heterogeneity in complete vaccination coverage. We perform a clustering analysis using spatial scan statistics to highlight geographical areas of lower-than-expected vaccination coverage and analyze the time-varying patterns of growth in vaccination coverage. We then fit partially pooled county-level logistic growth models to observed vaccination rates over time to better understand vaccination trajectories and progress across the country.

### US county-level vaccination data vary in quality

Comparison of the state-reported vaccination data and CDC-reported data shows large discrepancies. In particular, the complete vaccination coverage for the counties in Texas, Georgia, West Virginia, Virginia, and Colorado, as well as some counties in New Mexico, California, Vermont, North Carolina, Minnesota and a number of other states are significantly underestimated in CDC reports (Figure S1). On the other hand, states tend to under-report vaccination in locations with large federally-serviced populations (i.e., military bases, Indian reservations, veterans, and incarcerated populations). We have integrated these two data sources to produces a more accurate understanding of the distribution of vaccine protection across the US at a fine spatial scale. (More information is available in Methods and Supplementary Figures).

Importantly we measure vaccination coverage for complete vaccine protection and account for the total population size of a county. In contrast to the varying metrics used by states, this metric provides a consistent numerator and denominator and so is directly comparable across counties. The numerator–complete vaccination coverage (i.e., one dose of a one-dose schedule or two doses of a two-dose schedule)–is the most epidemiologically-relevant metric in the context of the Delta variant, which severely impairs partial vaccine protection [34]. Likewise, the denominator considers the entire susceptible population, not just those eligible for vaccination, which is the most informative metric for infectious disease transmission. These choices in measurement have important implications for public health; for example, the state of Vermont has been touted as having carried out the most successful COVID-19 vaccination campaign and reached a target of “80% coverage” on June 14, 2021 [35]. However, this target was only for partial vaccination coverage in individuals aged 12 years and above, and as of August 1, 2021, only 43-65% of the total populations have complete vaccine protection in Vermont counties.

### COVID-19 vaccination in the US demonstrates high spatial heterogeneity

The resulting distribution of COVID-19 vaccination at the US county scale shows significant geographic variability (Figure 1). By the week of August 1, 2021 (week 30), US counties vary from 9% to 90% complete vaccination protection against COVID-19. Additionally, there is significant variation in vaccination coverage across counties within a state, particularly in the western US (Figure S2), emphasizing the need to characterize vaccination at a fine scale. We also find significant spatial autocorrelation (Moran’s I = 0.57 during week 30) in vaccination distribution (Supplementary Figure S3).

**Figure 1:**
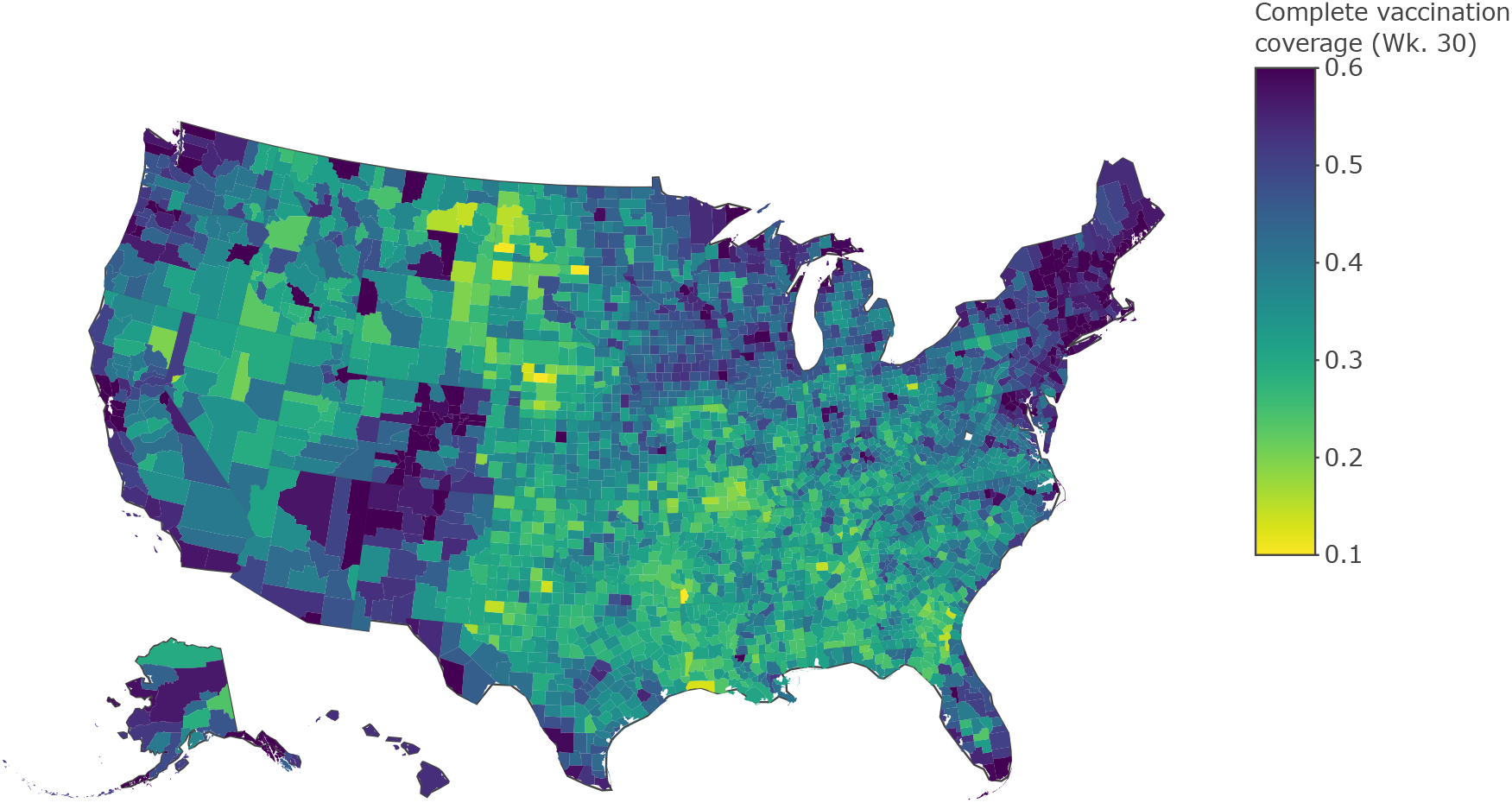
The COVID-19 county-level vaccination landscape. We show complete vaccination coverage for each US county using our integrated data set for the week ending Aug 1, 2021. There is significant heterogeneity across the country and within each state. Coverage levels vary from 9% to 90%, with a mean of 39%.

To identify vulnerable regions of lower than expected vaccination, we use the spatial scan statistic to identify spatial clusters of undervaccination. As of August 1, 2021, a total of 146 spatial clusters with fewer vaccination cases than expected are detected. We provide the top five clusters, ordered by their likelihood ratios (Figure 2 and Table S3), all of which have p-values < 1 × 10^−17^. Cluster 1 is found in eastern Texas/western Louisiana, Cluster 2 is found in eastern Alabama/western Georgia, Cluster 3 is found in northern Arkansas/southern Missouri, Cluster 4 is found in northern Texas/eastern New Mexico/southwestern Oklahoma, and Cluster 5 is found in northern Mississippi/northern Alabama/southwestern Tennessee. Moreover, these clusters are all found in the southern US, have populations of at least 2.2 million people, and are made up primarily of rural counties (with populations smaller than 10,000 individuals).

**Figure 2:**
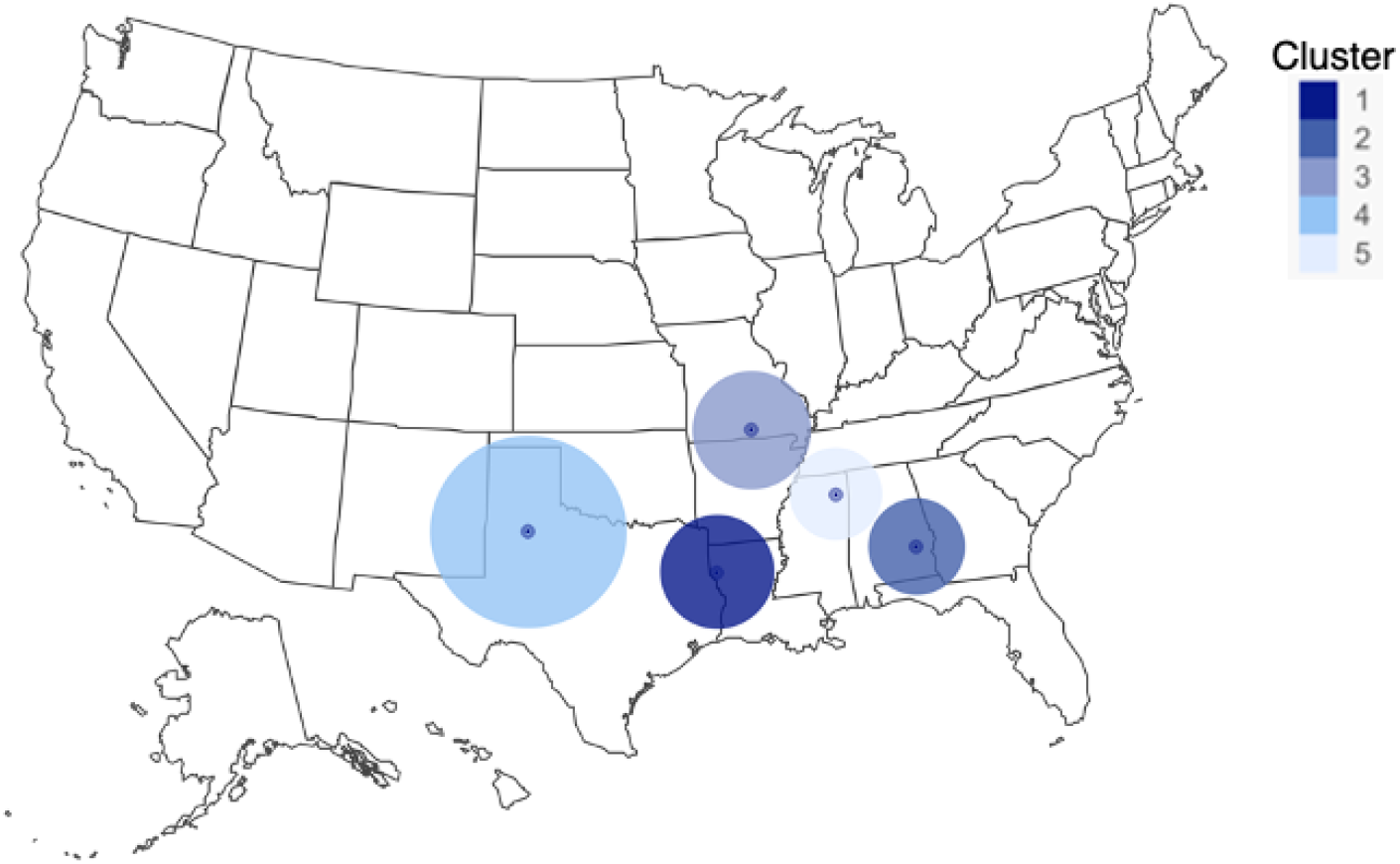
Clusters of COVID-19 undervaccination. The five spatial clusters with the highest likelihood of having lower than expected COVID-19 vaccination rates are concentrated in the southern US. Clusters span counties primarily in Louisiana, Texas, Alabama, Mississippi, Arkansas, and Missouri, but do not respect state borders.

Repeated analyses on the first complete week of each month from May-August yield a similar geographical distribution in clusters of undervaccination (Figure S4).

### US COVID-19 vaccination patterns are characterized by surges and stalls

County-level vaccination progress shows significant variation between and within states (Figure 3). All counties exhibit a similar sigmoid-shaped trajectory, but vary in their initial vaccine uptake rates, the time it takes for the vaccination campaign to decelerate, and the rate at which uptake stabilizes. Notably, while vaccination uptake has slowed throughout the country, vaccination coverage remains far from reaching 100% of the eligible population (12+ years of age) in most communities.

**Figure 3:**
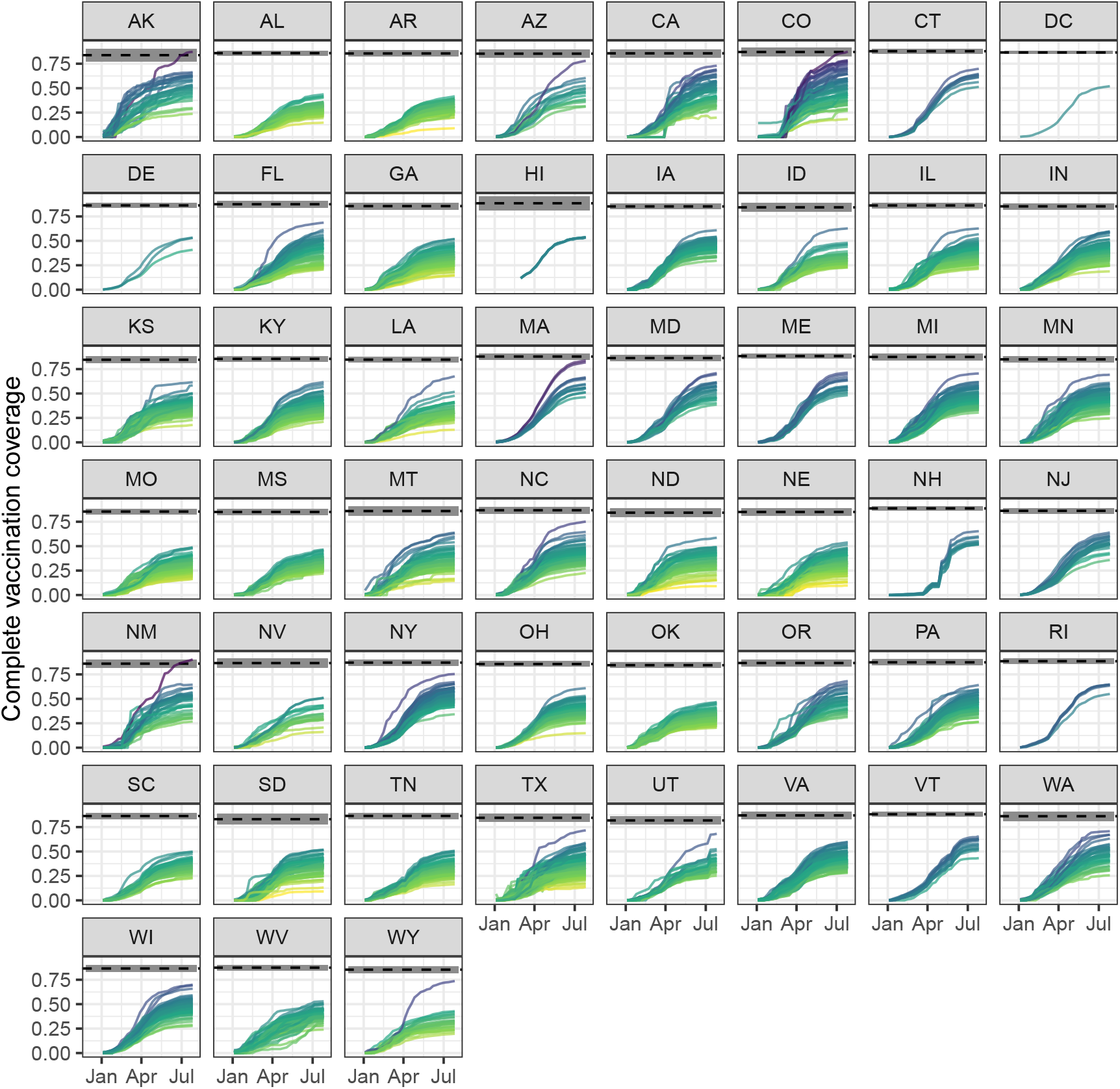
COVID-19 vaccination progress over time. Vaccination progress in each county generally follows a sigmoid shape through time, with an initial period of accelerated growth followed by a deceleration. Vaccination growth rate in most counties slows after June 2021, despite low coverage levels. Counties are colored by the complete vaccination coverage they reach by week 30, with lighter shades denoting lower coverage and darker shades denoting higher coverage. The average eligible population size (aged 12 years and above) for each state is denoted by the dashed black line, with a range of one standard deviation shaded in gray.

The stagnation we observe in COVID-19 vaccination rates represents a lost opportunity to increase vaccination immunity in populations, particularly in the face of recent surges in cases spurred on by the Delta variant [4]. The majority of US COVID-19 cases were caused by the Delta variant after week 25 of 2021 [36]. Thus, we consider the period preceding this time (week 21, ending May 30, to week 25, ending June 27) to identify counties that stalled in their vaccination efforts. Nearly half of US counties saw some period of stalling vaccination rates during this period, with longer periods of low growth concentrating in the South and the Plains area of the country (Figure 4a).

**Figure 4:**
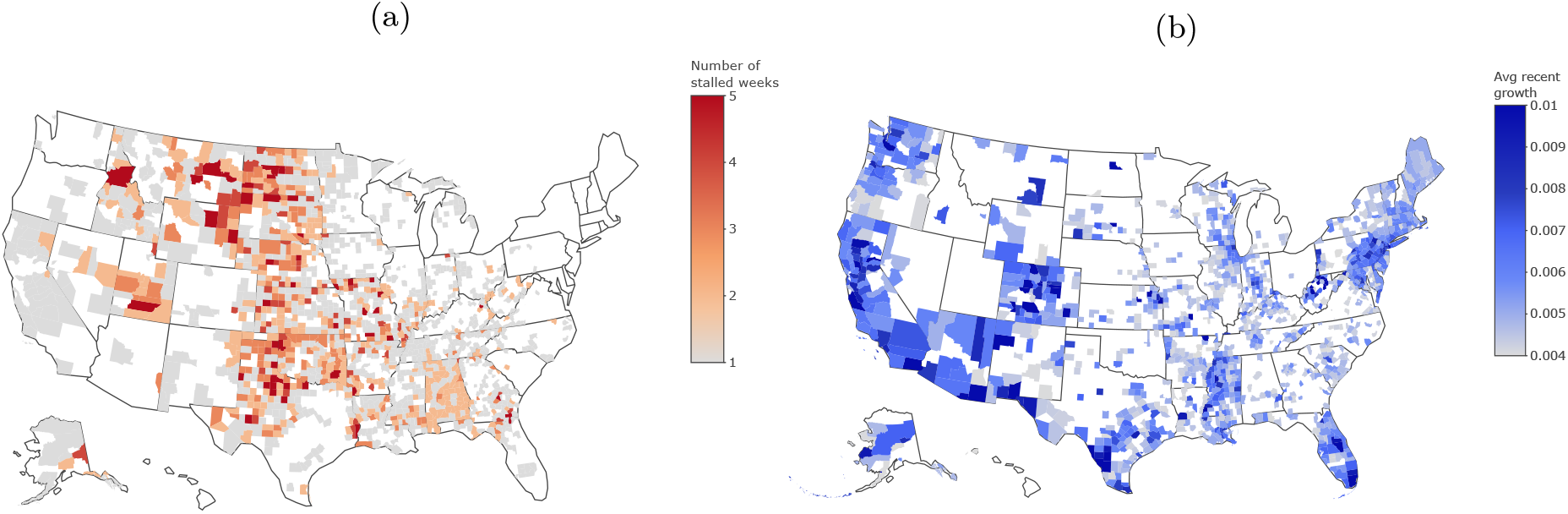
The missed opportunities and efforts to catch up on variant transmission mitigation. (a) In the weeks preceding the predominance of the Delta variant (week 21 to week 25), almost 50% of US counties experienced stalling in vaccination progress. Counties in the Plains exhibit longer periods of stalling. (b) With widespread circulation of the Delta variant (week 26 to week 30), the average vaccination growth rate is significant in approximately a third of US counties, but not in many of the previously stalled communities. (We note that Utah is omitted from panel (b) due to data issues introduced during weeks 28-30.)

We then examine the impact of increasing Delta variant prevalence in July on vaccination rates [36]. Because the Delta variant has increased the risk of infection and subsequent hospitalization (see [37, 38]), we consider the hypothesis that vaccination rates would increase in response to the increased risk [39]. We consider the county-level average growth rate of vaccination from the week of July 4, 2021 (week 26) to the week of August 1, 2021 (week 30). We find that average growth rate during this period is elevated in about a third of counties (Figure 4b). These counties are heterogeneously distributed throughout the country, largely clustered along the east and west coasts, and tend to have larger populations (with the average population of growth counties being more than twice as large as the average US county population). Importantly, when compared with the counties in which vaccination rates stalled, the two groups are largely non-overlapping. Indeed, the growth rate in the stalled counties during weeks 26-30 is only 0.0033, on average, meaning that vaccination rates in these communities continue to stall despite increased Delta transmission.

Lastly, we fit the observed data on vaccination over time from week 1 to week 25 to a simple model of growth and use comparison with this model to identify unusual vaccination dynamics post-week 25. In particular, we fit county-level vaccination coverage time series to models of logistic growth. We find that the model characterizes the observed data well (Figure S6 & Figure S7), with a first stage of rapid growth in vaccination, separated by an inflection point (Figure S8) from a second stage during which growth in vaccination slows. Rather than interpret model coefficients directly, we treat the logistic growth models as null models and examine deviation of observed county-level vaccination rates from these null models. In particular, we focus on deviations from modeled expectations during the second stage of the US vaccination campaign (i.e., week 15 and beyond). Based on these deviations, we find that counties may be grouped into four classes of dynamics (Figure 5 & Figure S9): a) counties that display strong adherence to our logistic growth model (e.g., Allegany County, MD). This is the most common outcome with 73% of all US counties in this class; b) counties in which the observed vaccination coverage begins to overshoot our model predictions in June or July of 2021 (e.g., Cochise County, AZ); c) counties in which growth rate is faster than expected during the second stage of vaccination, overshooting model expectations (e.g., Durham County, NC); and d) counties in which vaccination grows rapidly in the first stage of the pandemic, and abruptly slows at the end, falling below expectations (e.g., Essex County, VT).

**Figure 5:**
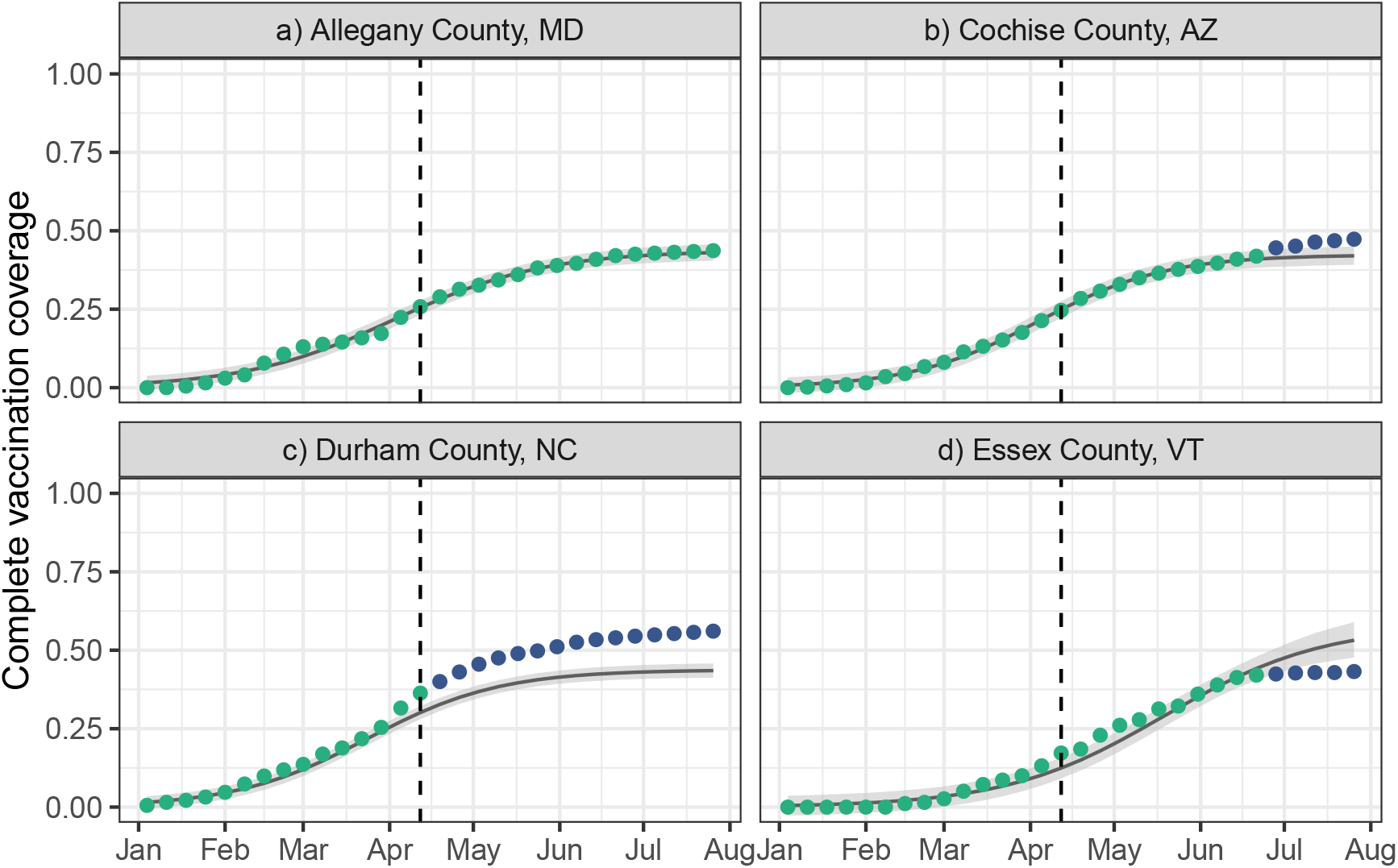
Vaccination dynamics compared to a simple model of growth. We provide examples of counties with the comparison of the vaccination data against the model fit from the logistic growth model. Weekly observations are colored by whether they fall inside (green) or outside (purple) the 95% prediction interval. The vertical dashed line marks week 15 (week ending April 18, 2021), the week after which deviations are identified. (a) The vaccination rates in Allegany County, MD show close adherence to the model expectation. All observations fall within the 95% prediction interval. (b) The vaccination rates in Cochise County, AZ rise above model expectations in late June. (c) The vaccination rates in Durham County, NC exceed model expectations beginning in mid-April. The vaccination rates in Essex County, VT are lower than model expectations starting in late June.

## Discussion

The US COVID-19 vaccination landscape has shaped the population health impacts of SARS-CoV-2 [9], structured the potential for local elimination alongside natural immunity and behavioral containment [1], and will drive the transition out of the pandemic to endemic circulation of SARS-CoV-2 [40]. However, the tracking of vaccination progress at a fine geographical scale and through time has not been a US public health priority.

Official vaccination data from public health agencies at the state and federal level are the gold standard for monitoring vaccination progress. However, we find that vaccination data reported by state health departments and the Centers for Disease Control and Prevention have discrepancies, and each captures different subsets of the total population. While states are responsible for allocating federally-allotted vaccination doses as well as tracking and reporting progress, the federal government is responsible for tracking vaccine administration within federally-serviced populations throughout the state. These separately serviced populations create the potential for states’ reported vaccination counts to be lower than those at the federal level as not all states integrate the data from these special populations. On the other hand, certain states do not share county-level vaccination data with the CDC due to privacy and other concerns (e.g., for all counties in Texas, and for small counties in Virginia, California and Massachusetts) [41]. These data issues impede efforts to accurately measure and track fine-scale vaccination progress and to identify target locations that may be undervaccinated. COVID-19 vaccination tracking is currently done through a network of immunization information systems designed to be a centralized data repository for vaccination information within each state. However, together they make up a disconnected patchwork of 64 software systems of varying data capacity and data quality [42]. Combined with significant inconsistencies in state policies on vaccination administration and data reporting, this has created a disordered data landscape which undermines planning for the next phase of the pandemic, and erodes an already crumbling public health data infrastructure in the US [43].

Although vaccination against COVID-19 has helped to dramatically lessen transmission rates in the US, cases have once again surged due to the more transmissible Delta variant [44, 4, 5]. The most affected states–in terms of daily case counts and hospitalizations–are found in the southern/southeastern US [4]. These hard hit states contain high-risk subpopulations with low vaccination coverage, as identified by our spatial clustering results. Importantly, the undervaccination clusters we identify make up a subset of communities within the states to which they belong, highlighting the need for fine-scale vaccination data and analysis to capture these vulnerabilities. Using metrics such as state averages obscures this county-level heterogeneity, yielding an inaccurate measure of local risk. Surges in these mostly rural areas run the risk of quickly overwhelming the local health care systems, which are often unequipped for a sudden influx of serious COVID-19 hospitalizations, and leading to increases in COVID-19-related mortality [44, 45]. Our clustering results also maintain a similar geographical pattern from May 2021 into August 2021 suggesting a persistence in low vaccination throughout the vaccination campaign. Notably, recent data estimate vaccine hesitancy to be limited to 10-20% of the population in these states (see e.g., [46]), suggesting that vaccine hesitancy explains only part of the remaining undervaccination. This result implies that there remains a substantial proportion of the unvaccinated population that is willing to be vaccinated but has not yet been able to do so; systemic issues of inaccessibility and inequality in healthcare are the likely culprit [47, 48].

Our results highlight that vaccination rates across the US significantly slowed during the period prior to the Delta variant’s predominance, resulting in a missed opportunity to prepare for the coming surge. In fact, public risk perception of COVID-19 appears to decrease and maintains its lowest levels during this period (Figure S5). In the weeks following the predominance of the Delta variant, vaccination rates have seen small increases, but these increases are heterogeneously distributed– rarely occurring in stalled counties with lower vaccination rates.

Our model-based fits to the observed vaccination data provide a reference with which to classify county-level vaccination dynamics. In particular, we hypothesize that vaccination growth dynamics follow logistic growth dynamics in which the vaccinated population grows in a positive acceleration stage while vaccine acceptance is high followed by a deceleration stage of increasing vaccine resistance. The structure of the vaccination phases in the US COVID-19 vaccination campaign prioritized individuals at high clinical risk and individuals in high transmission-risk occupations, followed by the general population [49]. We find that the inflection point (the transition between the two stages of growth) in our model fits corresponds to the shift in vaccination from the high-priority populations to the general population around mid-April (Figure S8), agreeing with our hypothesis.

We find that most counties are highly consistent with the model dynamics and focus on the locations in which the observed dynamics deviate from model expectations to further understand vaccination progress. One group of communities surges in vaccination in June after reaching a plateau previously. For these locations, we speculate that the closing of mass vaccination sites in June resulted in a shift of resources towards community vaccination and mobile vaccination sites and the increased access may be responsible for this growth [50]. Another set of counties surges in vaccination in July after previously stalling. We hypothesize that this increase is driven by an increase in COVID-19 risk perception following reports of increased Delta transmission around this time (Figure S5). Other communities, primarily in North Carolina, saw a larger surge in the second stage of vaccination than model expectations. While North Carolina largely adhered to the guidelines of the Advisory Committee on Immunization Practices [51] for vaccine distribution, it did not prioritize adults with high-risk medical conditions or adults over 60 years of age [49]. Thus we hypothesize that this difference in vaccine prioritization led to these vulnerable groups urgently getting vaccinated at the first opportunity, producing a large deviation from the expected dynamics around week 14 (which then persists given the cumulative nature of the data). The last group of communities, primarily in Vermont, plateaus at a lower vaccination rate than the model expectations. This behavior could be due to a more rapid acceleration during the first stage of vaccination than in other states, owing to a strong community effort to reach vaccination goals. Alternatively, we suggest that reaching targets set by the state (e.g., 80% partial vaccination coverage in the 12+ population–which was reached on June 14, 2021 [35])–may have dampened the momentum of the ongoing campaign.

Despite the influence of vaccination on SARS-CoV-2 transmission dynamics and associated morbidity, spatiotemporal heterogeneity in COVID-19 vaccination in the US has never before been systematically characterized. Indeed, systematic issues in data quality have impaired earlier analyses: ignoring federally-serviced populations, missing small counties, or underestimating coverage in entire states. The vaccination heterogeneity we identify not only increases local disease transmission, but also elevates risk across entire geographic regions; persistent clusters of undervaccination in the southeastern United States lead to increased transmission all over the country–including break-through infections in vaccinated individuals. However, undervaccination cannot be solely attributed to vaccine hesitancy. Systematic healthcare inequality has deprived many of the protection afforded by vaccination against COVID-19, and undervaccination continues to persist in the most vulnerable parts of the nation, despite recent surges of transmission and mortality. A sustained commitment to increasing vaccination–including expanded vaccine access and vaccine mandates–are necessary steps on a path toward local suppression of COVID-19. Significant investment in strengthening the US public health data infrastructure is urgently needed to handle the next public health crisis.

## Data Availability

Data are available at https://doi.org/10.7910/DVN/BFRIKI

## Acknowledgments

Author affiliations: Department of Biology, Georgetown University, Washington, District of Columbia, United States.

AT performed all analyses, interpreted the results, and drafted the manuscript. ZS performed the model analyses, interpreted results, and edited the manuscript. AM collected the data, interpreted the results, and edited the manuscript. SB designed the study, collected the data, guided the analysis, interpreted the results, and edited the manuscript. All authors read and approved the final manuscript.

This work was supported by the National Institute of General Medical Sciences of the National Institutes of Health under award number R01GM123007. The content is solely the responsibility of the authors and does not necessarily represent the official views of the National Institutes of Health.

Data are available at https://doi.org/10.7910/DVN/BFRIKI.

Preprint available at https://doi.org/10.1101/2021.10.04.21263345. The authors declare no competing interests.

CDC Centers for Disease Control and Prevention

## Supplementary Information

### Supplementary Methods

### Data collation and cleaning

We choose complete vaccination data over partial vaccination data because the latter have more gaps, making analysis challenging. We also note that our county-level vaccination estimates only capture those records for which county residence information has been provided and the vaccinated individuals live in the state that they are vaccinated in. For states which provide the proportion of records without county residence information or out of state, we find that these uncounted vaccinations make up less than 10% of all vaccinations in most states. (See Supplementary Table S1 for details.)

When our integration of the CDC-reported and state-reported data across time results in discontinuities in the data due to a discrepancy between the two data sources, we scale vaccination coverage for the earlier time period so it is continuous with the later data.

### Logistic growth model

The Bayesian logistic growth model is defined as:

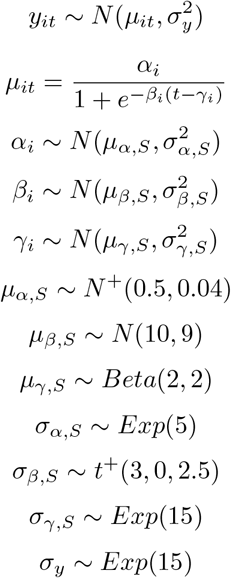

where *N* ^+^ and *t*^+^ refer respectively to normal and student-t distributions truncated at 0 to maintain positive real support only.

We fit these Bayesian models in Stan with brms version 2.15.0 [52, 53] and the cmdstanr version 0.4.0.9000 backend [54] using Hamiltonian Monte Carlo with 4 chains. For each chain, we use 5,000 warmup iterations and 5,000 sampling iterations, for a total of 20000 post-warmup samples. For each model, 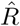 statistics are less than or equal to 1.01 and there were no divergences reported, suggesting unbiased numerical sampling and convergence between chains.

### Supplementary Figures

**Figure S1:**
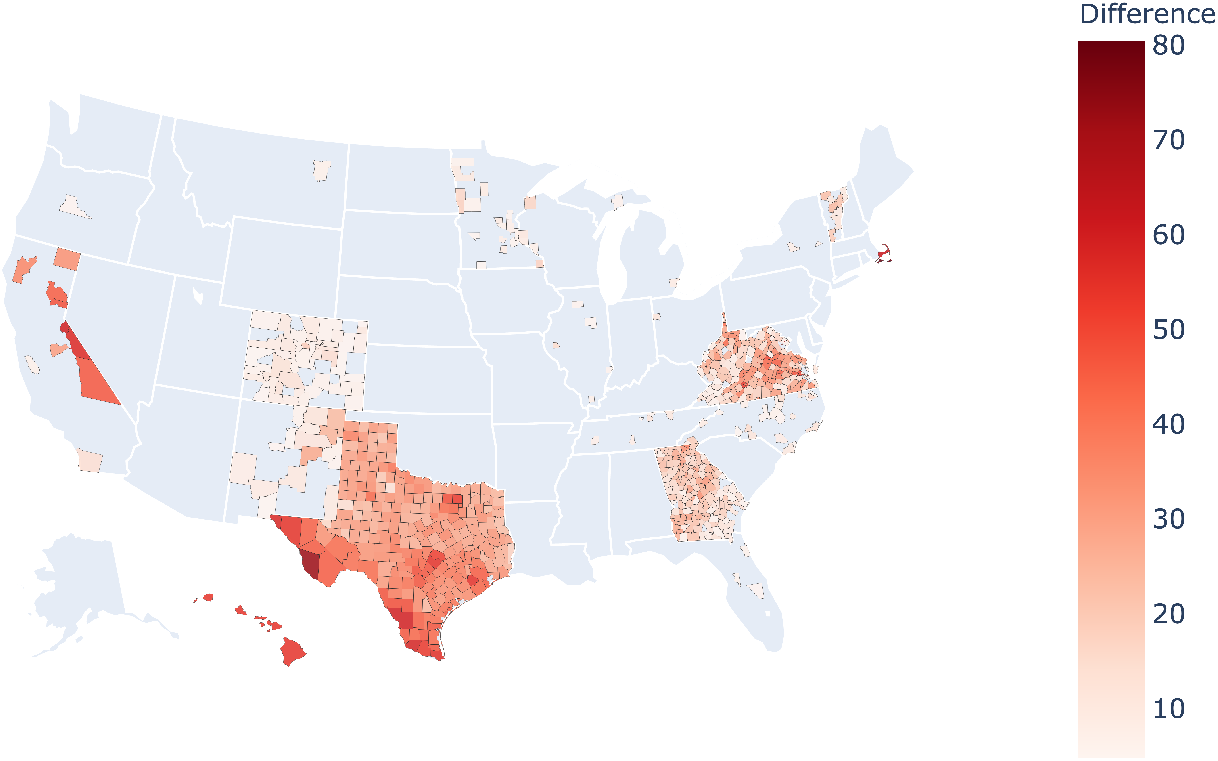
Incomplete CDC-reported vaccination data: We compare CDC-reported data to state-reported vaccination data for week 30, and identify the states for which the CDC data is incomplete. The map shows the difference between the complete vaccination coverage reported by the state and that reported by the CDC.

**Figure S2:**
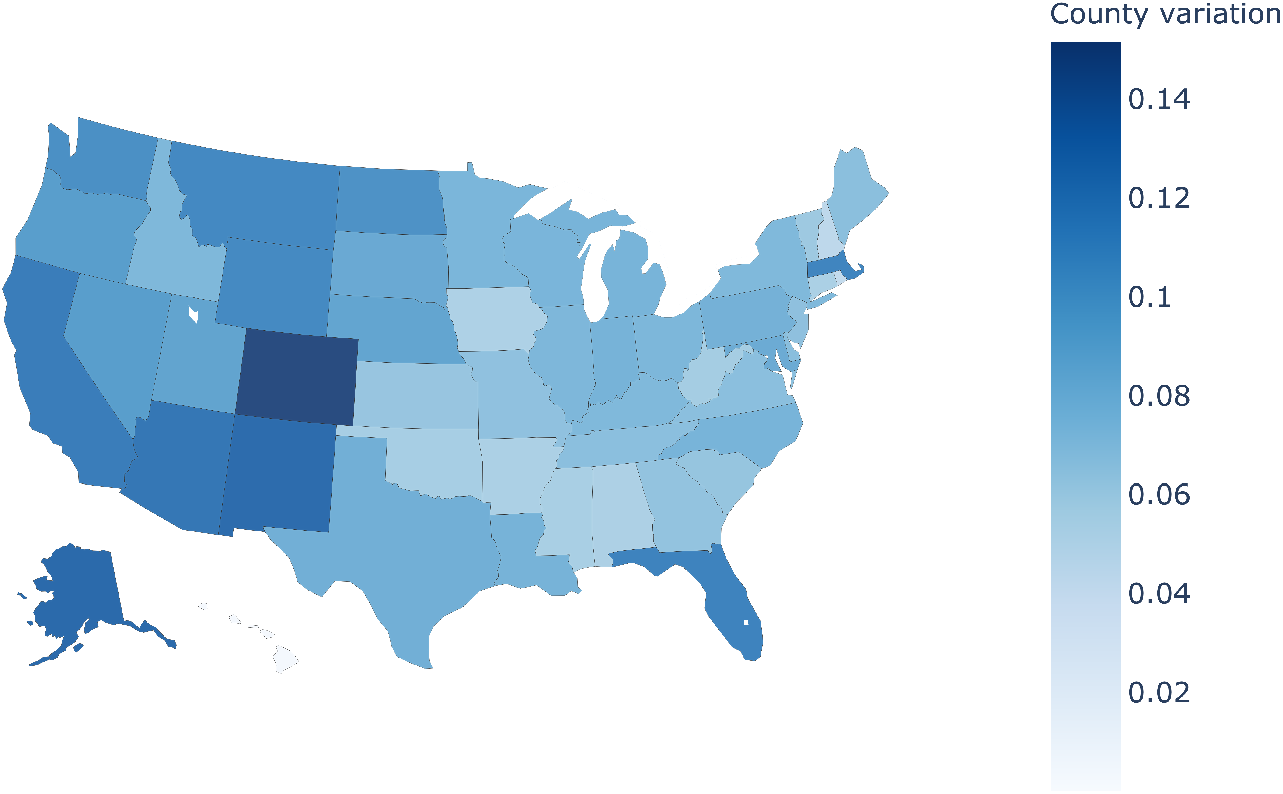
County-level variation in vaccination: For each state, we measure the standard deviation in vaccination coverage at week 30 across all counties within that state

**Figure S3:**
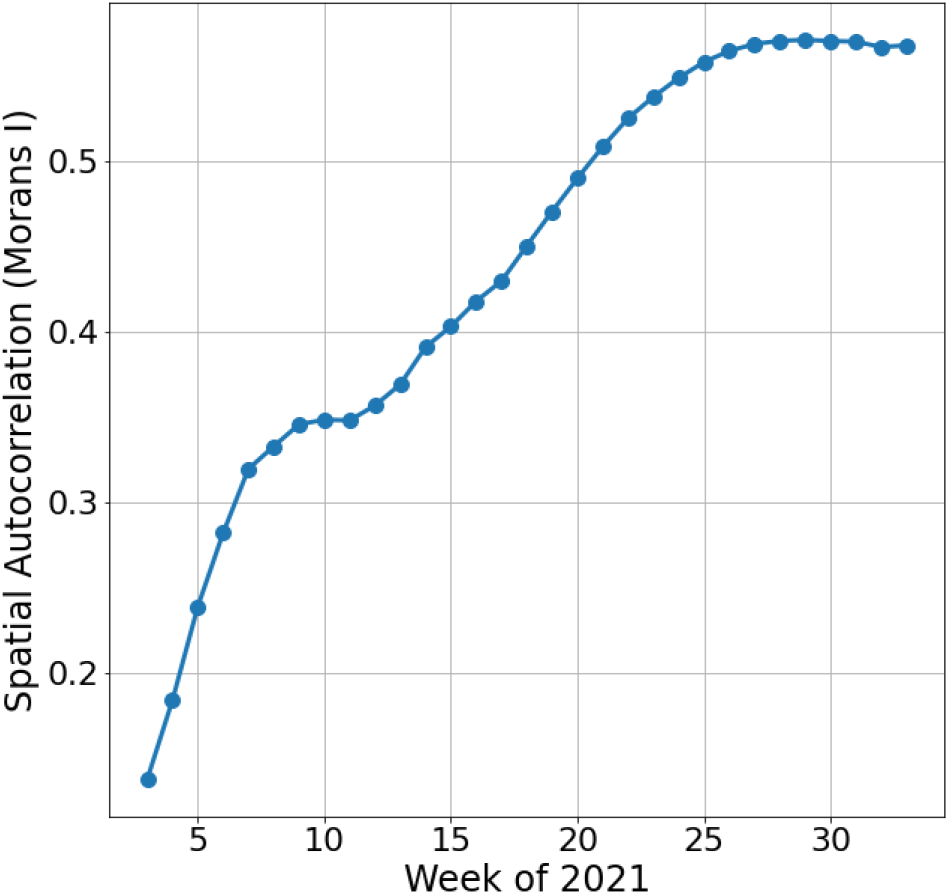
Spatial autocorrelation in county-level complete vaccination coverage as measured by Moran’s I for every week of data through 2021.

**Figure S4:**
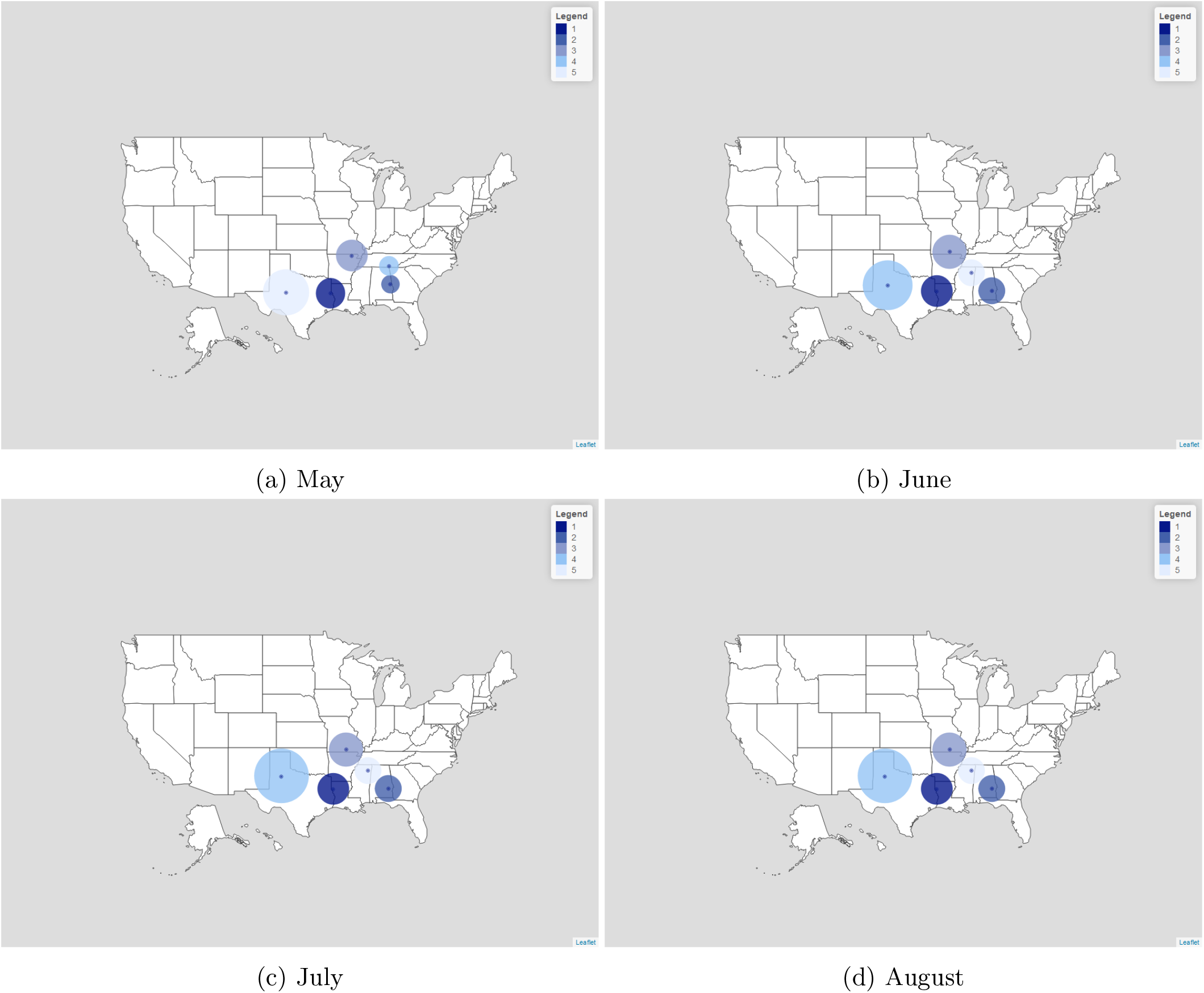
Persistence of spatial clusters across time. The most likely clusters of less-than-expected COVID-19 vaccination are found in the same general area across time from May to August of 2021. Specifically, we perform spatial clustering on the first week of each of these months.

**Figure S5:**
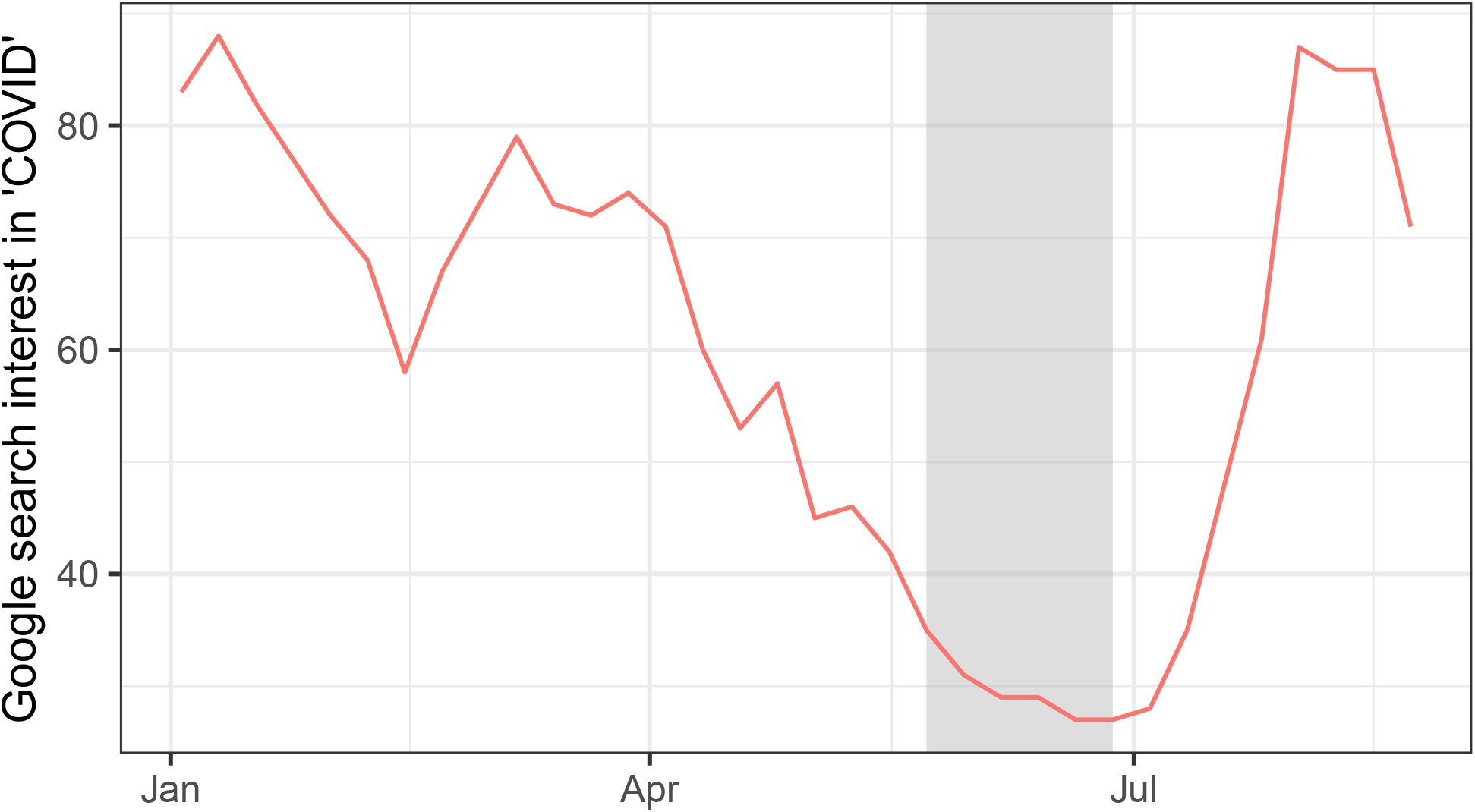
Evolving COVID risk perception through 2021: We evaluate information-seeking behavior based on Google Trends searches for the term “COVID” as a measure of COVID-19 risk perception. Values for search interest have been normalized so that a value of 100 denotes peak popularity for a given search term. The light gray, shaded area represents the period from week 21 to week 25.

**Figure S6:**
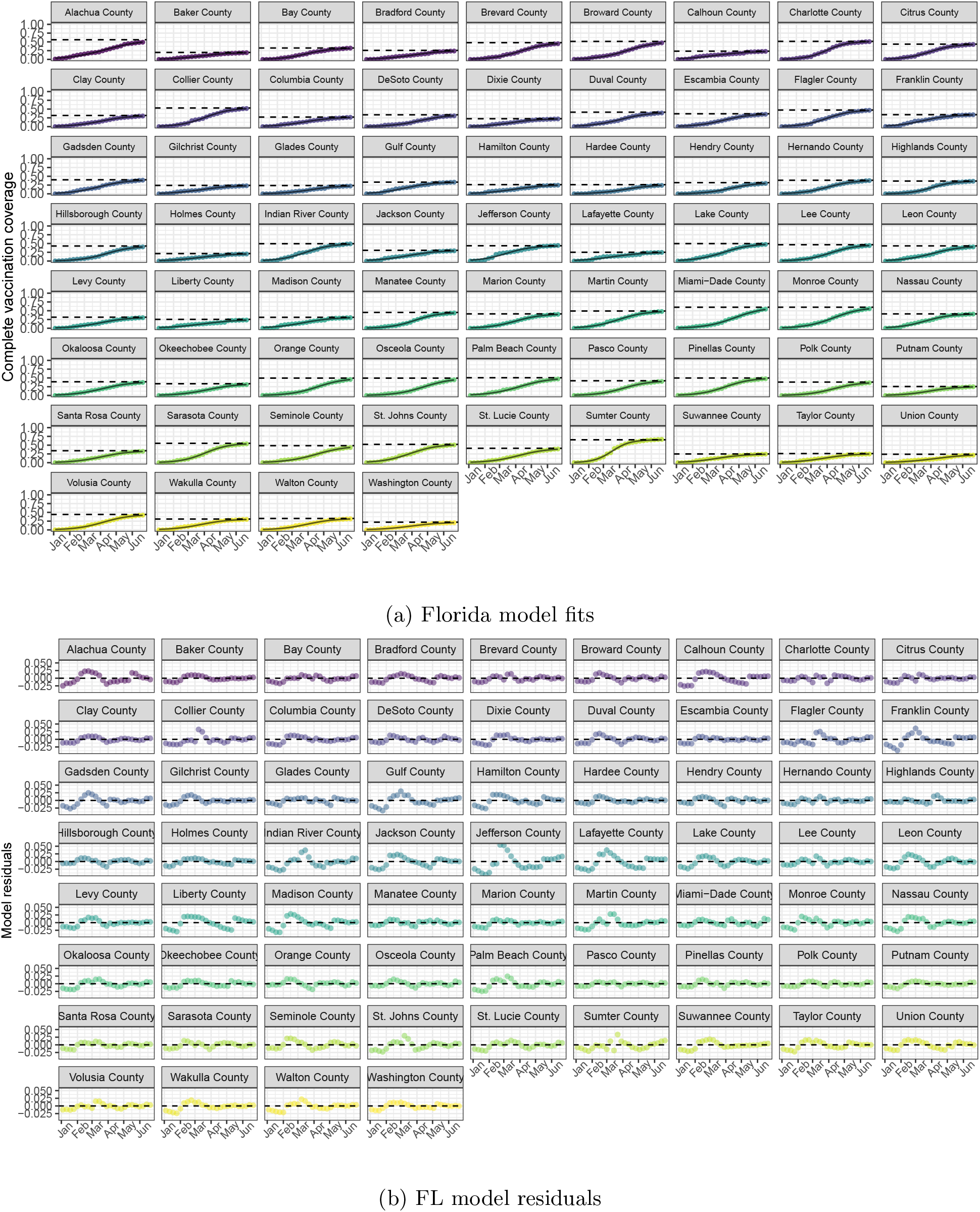
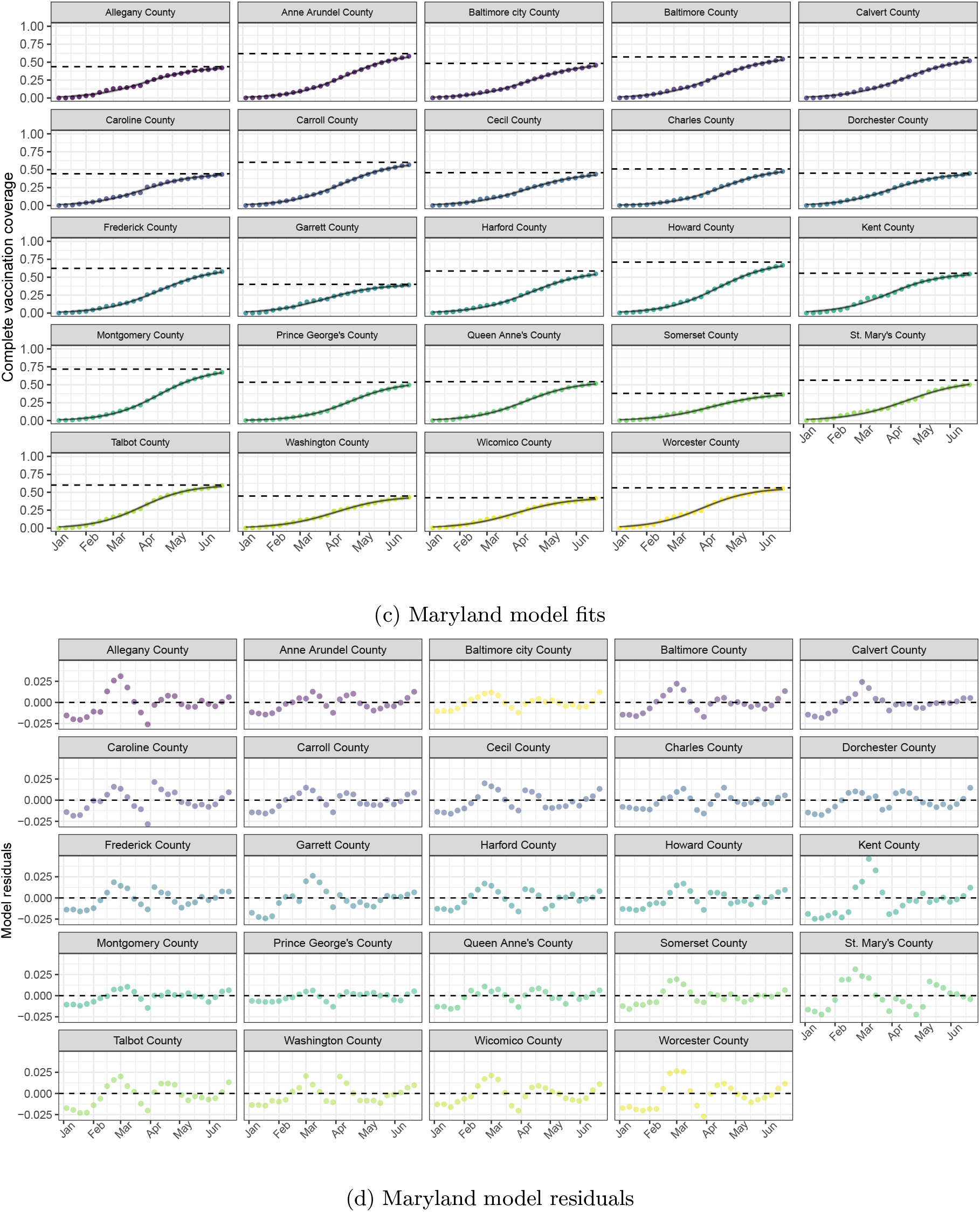
Selected model fits and residuals. We provide here the model fits and raw residuals for (a & b) Florida and (c & d) Maryland, respectively, within the modeled period (the week of January 10, 2021 to the week of June 27, 2021). In each state’s pair of panels, observed data are compared with the model estimates (solid black line), with the 95% prediction interval shaded in light gray. The dashed horizontal line represents the model-estimated asymptote for each county. Here, the observed vaccination coverage data over time closely follow our model predictions.

**Figure S7:**
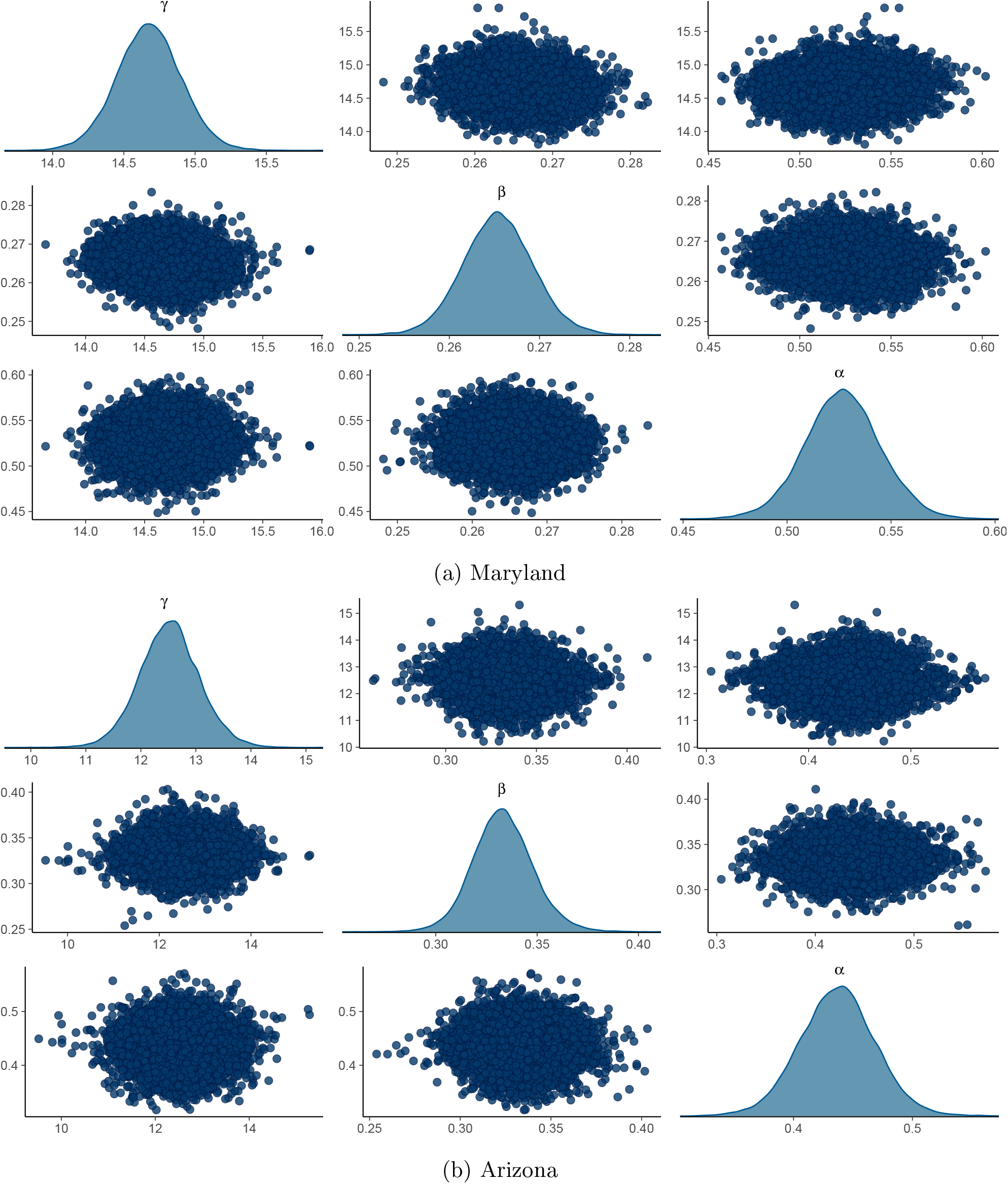
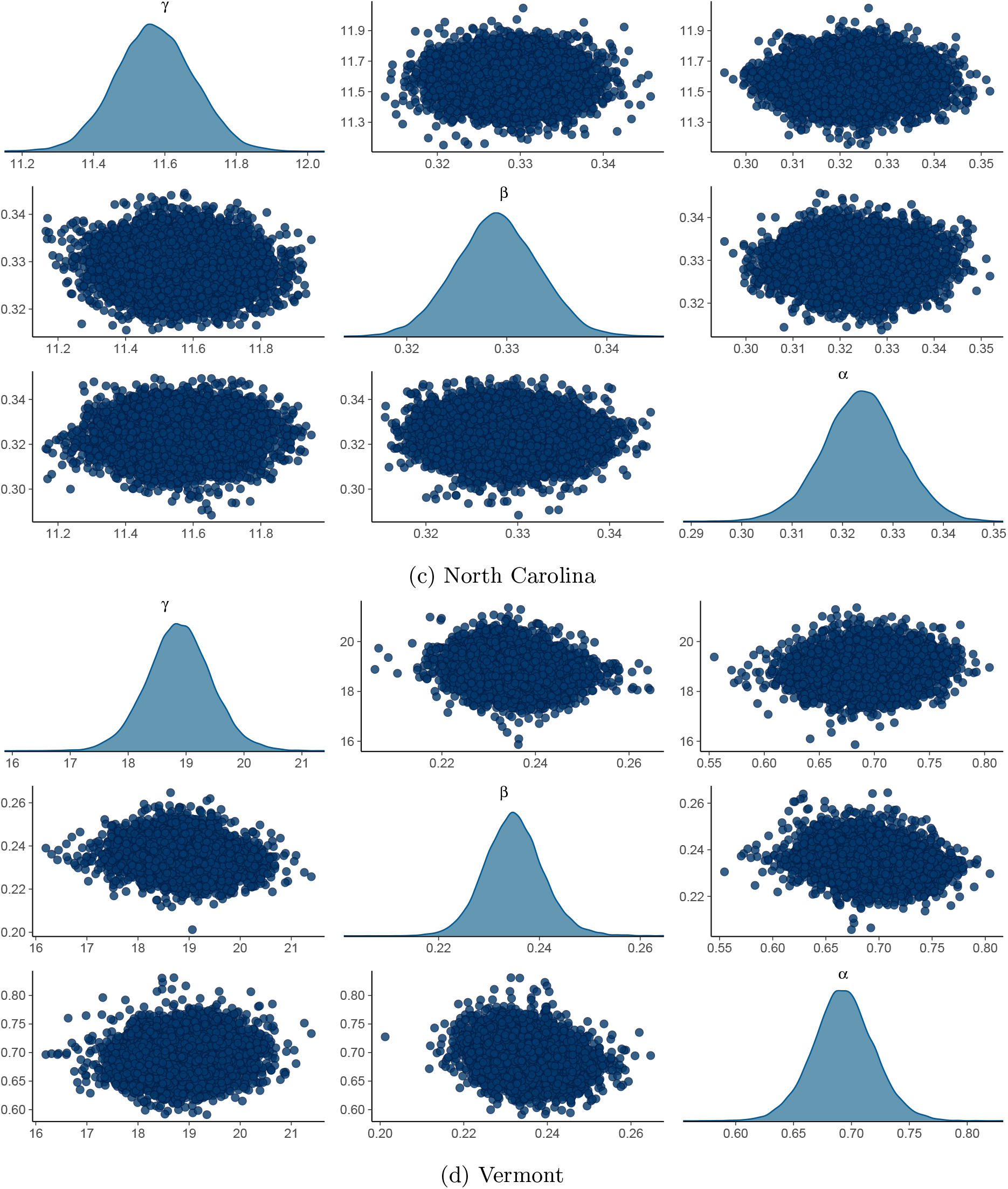
Density plots for marginal posterior distributions of main model parameters. We provide here a selection of marginal distributions for the four state models referenced in Figure 5. *α* represents the asymptote parameter, *β* represents the rate parameter, and *γ* represents the inflection point parameter. Univariate marginal distributions are along the diagonal. Bivariate distributions are represented by the off-diagonal scatterplots.

**Figure S8:**
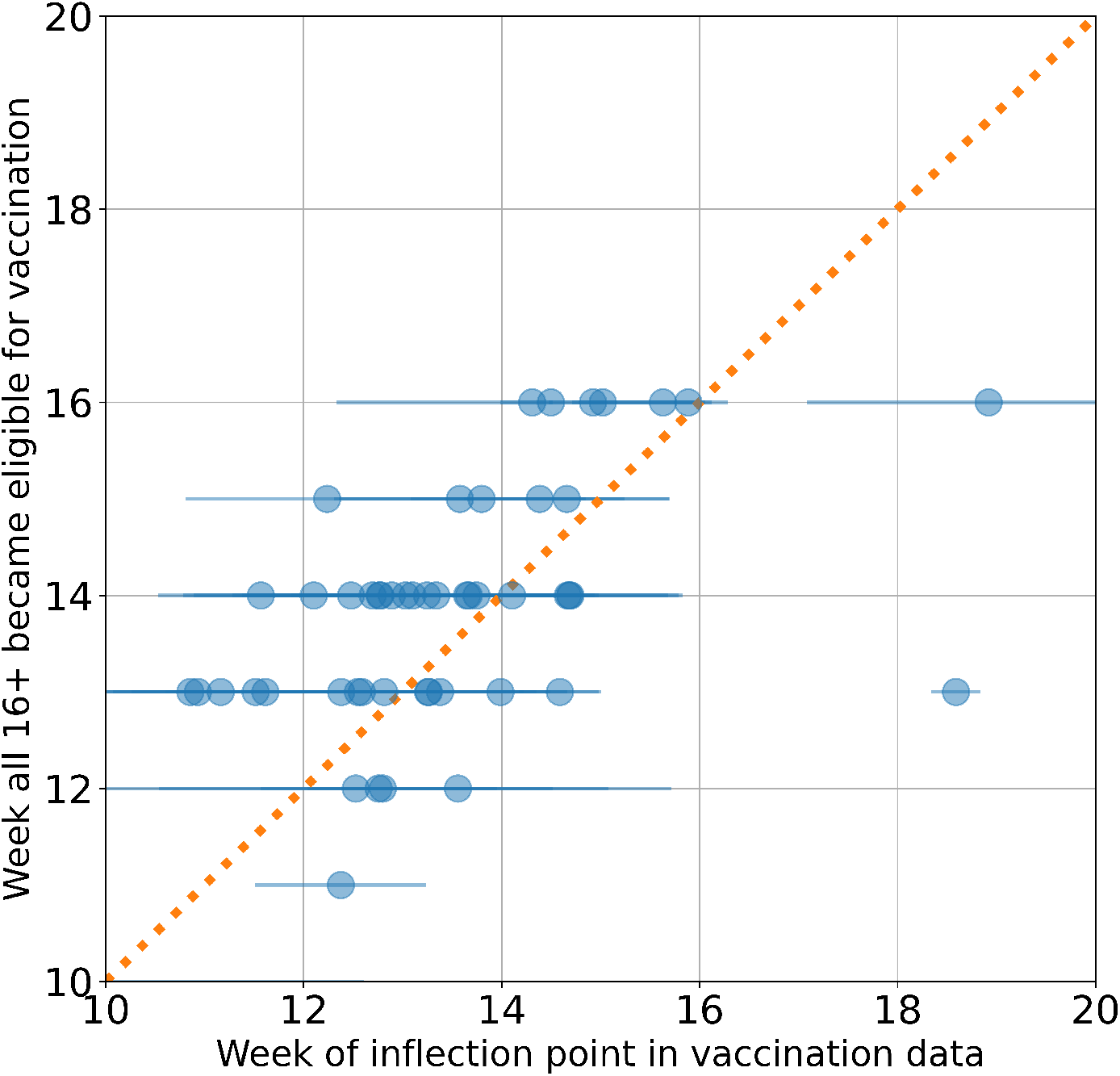
Transitions in vaccination We show the relationship between [x-axis] the model-fitted timing (week of 2021) of the inflection point (i.e., the transition between acceleration and deceleration in vaccination progress) for each state, with the standard deviation across counties and [y-axis] the week at which the state opened vaccination eligibility to the general public. The two outliers are NH and VT and NH has a significant discontinuity in its data making the model fit less accurate.

**Figure S9:**
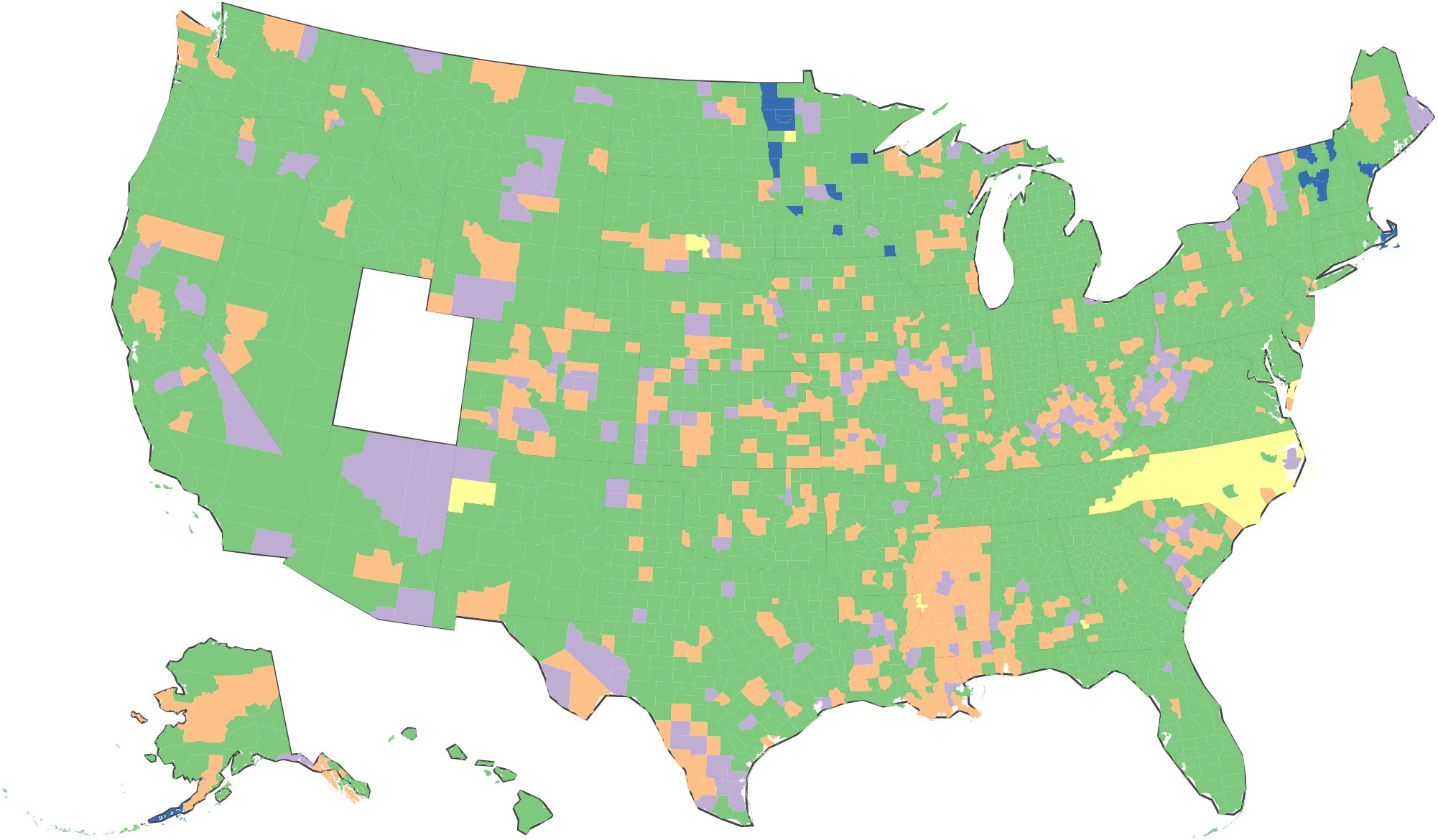
US counties may be grouped into four classes based on model deviations. We group counties by how they deviate from our logistic growth model–representing our expectations. Counties in green show close adherence to our models and represent the majority of cases. Counties in lavender overshoot our model predictions beginning in June 2021, while counties in orange do so beginning in July 2021. Counties in yellow overshoot our model predictions as early as April 2021. Counties in blue undershoot our model predictions. Note: Utah is omitted from this map due to structural issues in the data from weeks 28 to 30 which would affect their grouping.

**Table S1:**
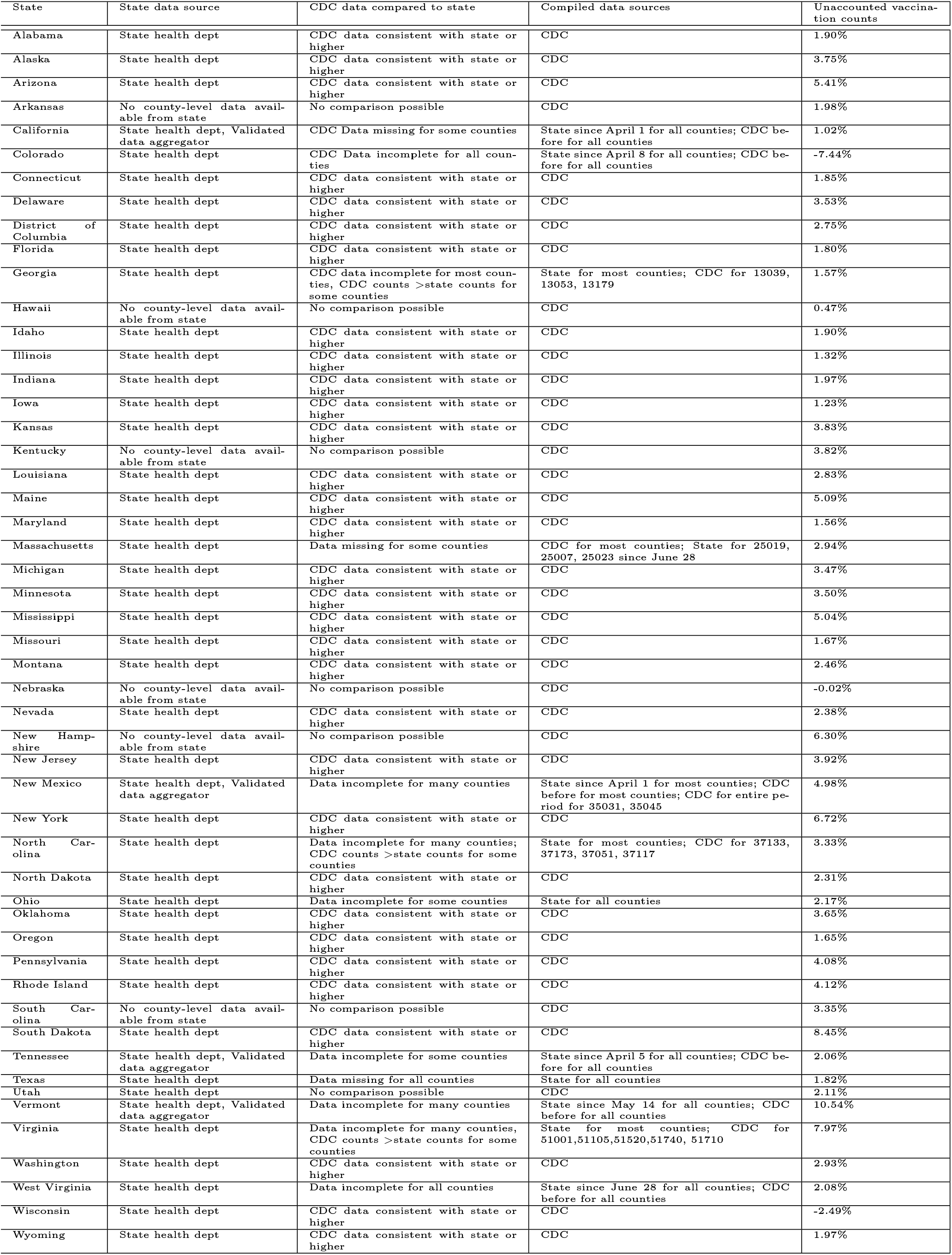
Details on the data source and consistency for each state. The unaccounted vaccination counts lists the vaccination counts (as a proportion of the state population size) for which we do not have county residence information.

| State | State data source | CDC data compared to state | Compiled data sources | Unaccounted vaccination counts |
| --- | --- | --- | --- | --- |
| Alabama | State health dept | CDC data consistent with state or higher | CDC | 1.90% |
| Alaska | State health dept | CDC data consistent with state or higher | CDC | 3.75% |
| Arizona | State health dept | CDC data consistent with state or higher | CDC | 5.41% |
| Arkansas | No county-level data available from state | No comparison possible | CDC | 1.98% |
| California | State health dept, Validated data aggregator | CDC Data missing for some counties | State since April 1 for all counties; CDC before for all counties | 1.02% |
| Colorado | State health dept | CDC Data incomplete for all counties | State since April 8 for all counties; CDC before for all counties | -7.44% |
| Connecticut | State health dept | CDC data consistent with state or higher | CDC | 1.85% |
| Delaware | State health dept | CDC data consistent with state or higher | CDC | 3.53% |
| District of Columbia | State health dept | CDC data consistent with state or higher | CDC | 2.75% |
| Florida | State health dept | CDC data consistent with state or higher | CDC | 1.80% |
| Georgia | State health dept | CDC data incomplete for most counties, CDC counts >state counts for some counties | State for most counties; CDC for 13039, 13053, 13179 | 1.57% |
| Hawaii | No county-level data available from state | No comparison possible | CDC | 0.47% |
| Idaho | State health dept | CDC data consistent with state or higher | CDC | 1.90% |
| Illinois | State health dept | CDC data consistent with state or higher | CDC | 1.32% |
| Indiana | State health dept | CDC data consistent with state or higher | CDC | 1.97% |
| Iowa | State health dept | CDC data consistent with state or higher | CDC | 1.23% |
| Kansas | State health dept | CDC data consistent with state or higher | CDC | 3.83% |
| Kentucky | No county-level data available from state | No comparison possible | CDC | 3.82% |
| Louisiana | State health dept | CDC data consistent with state or higher | CDC | 2.83% |
| Maine | State health dept | CDC data consistent with state or higher | CDC | 5.09% |
| Maryland | State health dept | CDC data consistent with state or higher | CDC | 1.56% |
| Massachusetts | State health dept | Data missing for some counties | CDC for most counties; State for 25019, 25007, 25023 since June 28 | 2.94% |
| Michigan | State health dept | CDC data consistent with state or higher | CDC | 3.47% |
| Minnesota | State health dept | CDC data consistent with state or higher | CDC | 3.50% |
| Mississippi | State health dept | CDC data consistent with state or higher | CDC | 5.04% |
| Missouri | State health dept | CDC data consistent with state or higher | CDC | 1.67% |
| Montana | State health dept | CDC data consistent with state or higher | CDC | 2.46% |
| Nebraska | No county-level data available from state | No comparison possible | CDC | -0.02% |
| Nevada | State health dept | CDC data consistent with state or higher | CDC | 2.38% |
| New Hampshire | No county-level data available from state | No comparison possible | CDC | 6.30% |
| New Jersey | State health dept | CDC data consistent with state or higher | CDC | 3.92% |
| New Mexico | State health dept, Validated data aggregator | Data incomplete for many counties | State since April 1 for most counties; CDC before for most counties; CDC for entire period for 35031, 35045 | 4.98% |
| New York | State health dept | CDC data consistent with state or higher | CDC | 6.72% |
| North Carolina | State health dept | Data incomplete for many counties; CDC counts >state counts for some counties | State for most counties; CDC for 37133, 37173, 37051, 37117 | 3.33% |
| North Dakota | State health dept | CDC data consistent with state or higher | CDC | 2.31% |
| Ohio | State health dept | Data incomplete for some counties | State for all counties | 2.17% |
| Oklahoma | State health dept | CDC data consistent with state or higher | CDC | 3.65% |
| Oregon | State health dept | CDC data consistent with state or higher | CDC | 1.65% |
| Pennsylvania | State health dept | CDC data consistent with state or higher | CDC | 4.08% |
| Rhode Island | State health dept | CDC data consistent with state or higher | CDC | 4.12% |
| South Carolina | No county-level data available from state | No comparison possible | CDC | 3.35% |
| South Dakota | State health dept | CDC data consistent with state or higher | CDC | 8.45% |
| Tennessee | State health dept, Validated data aggregator | Data incomplete for some counties | State since April 5 for all counties; CDC before for all counties | 2.06% |
| Texas | State health dept | Data missing for all counties | State for all counties | 1.82% |
| Utah | State health dept | No comparison possible | CDC | 2.11% |
| Vermont | State health dept, Validated data aggregator | Data incomplete for many counties | State since May 14 for all counties; CDC before for all counties | 10.54% |
| Virginia | State health dept | Data incomplete for many counties, CDC counts >state counts for some counties | State for most counties; CDC for 51001,51105,51520,51740, 51710 | 7.97% |
| Washington | State health dept | CDC data consistent with state or higher | CDC | 2.93% |
| West Virginia | State health dept | Data incomplete for all counties | State since June 28 for all counties; CDC before for all counties | 2.08% |
| Wisconsin | State health dept | CDC data consistent with state or higher | CDC | -2.49% |
| Wyoming | State health dept | CDC data consistent with state or higher | CDC | 1.97% |

**Table S2:**
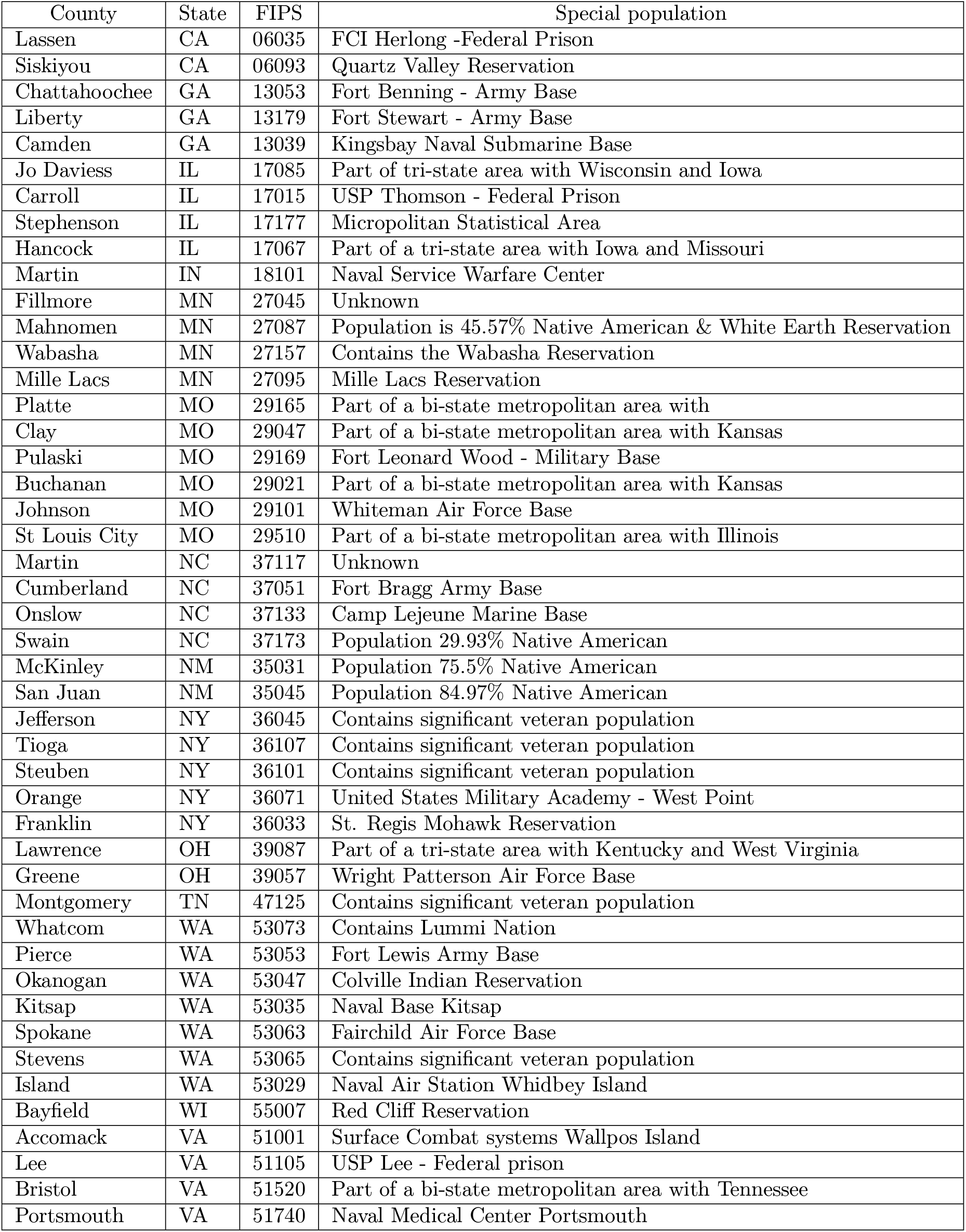
Counties for which special populations (such as Native American reservation populations, military personnel, veterans, and incarcerated populations) are not counted by the state but are included in vaccination counts reported by the CDC. We also include counties that are split between multiple states on this list; Additionally there are two counties for which we have not yet identified a reason for their underestimation in the state data; however we still include the CDC data for these locations.

| County | State | FIPS | Special population |
| --- | --- | --- | --- |
| Lassen | CA | 06035 | FCI Herlong -Federal Prison |
| Siskiyou | CA | 06093 | Quartz Valley Reservation |
| Chattahoochee | GA | 13053 | Fort Benning - Army Base |
| Liberty | GA | 13179 | Fort Stewart - Army Base |
| Camden | GA | 13039 | Kingsbay Naval Submarine Base |
| Jo Daviess | IL | 17085 | Part of tri-state area with Wisconsin and Iowa |
| Carroll | IL | 17015 | USP Thomson - Federal Prison |
| Stephenson | IL | 17177 | Micropolitan Statistical Area |
| Hancock | IL | 17067 | Part of a tri-state area with Iowa and Missouri |
| Martin | IN | 18101 | Naval Service Warfare Center |
| Fillmore | MN | 27045 | Unknown |
| Mahnomen | MN | 27087 | Population is 45.57% Native American & White Earth Reservation |
| Wabasha | MN | 27157 | Contains the Wabasha Reservation |
| Mille Lacs | MN | 27095 | Mille Lacs Reservation |
| Platte | MO | 29165 | Part of a bi-state metropolitan area with |
| Clay | MO | 29047 | Part of a bi-state metropolitan area with Kansas |
| Pulaski | MO | 29169 | Fort Leonard Wood - Military Base |
| Buchanan | MO | 29021 | Part of a bi-state metropolitan area with Kansas |
| Johnson | MO | 29101 | Whiteman Air Force Base |
| St Louis City | MO | 29510 | Part of a bi-state metropolitan area with Illinois |
| Martin | NC | 37117 | Unknown |
| Cumberland | NC | 37051 | Fort Bragg Army Base |
| Onslow | NC | 37133 | Camp Lejeune Marine Base |
| Swain | NC | 37173 | Population 29.93% Native American |
| McKinley | NM | 35031 | Population 75.5% Native American |
| San Juan | NM | 35045 | Population 84.97% Native American |
| Jefferson | NY | 36045 | Contains significant veteran population |
| Tioga | NY | 36107 | Contains significant veteran population |
| Steuben | NY | 36101 | Contains significant veteran population |
| Orange | NY | 36071 | United States Military Academy - West Point |
| Franklin | NY | 36033 | St. Regis Mohawk Reservation |
| Lawrence | OH | 39087 | Part of a tri-state area with Kentucky and West Virginia |
| Greene | OH | 39057 | Wright Patterson Air Force Base |
| Montgomery | TN | 47125 | Contains significant veteran population |
| Whatcom | WA | 53073 | Contains Lummi Nation |
| Pierce | WA | 53053 | Fort Lewis Army Base |
| Okanogan | WA | 53047 | Colville Indian Reservation |
| Kitsap | WA | 53035 | Naval Base Kitsap |
| Spokane | WA | 53063 | Fairchild Air Force Base |
| Stevens | WA | 53065 | Contains significant veteran population |
| Island | WA | 53029 | Naval Air Station Whidbey Island |
| Bayfield | WI | 55007 | Red Cliff Reservation |
| Accomack | VA | 51001 | Surface Combat systems Wallpos Island |
| Lee | VA | 51105 | USP Lee - Federal prison |
| Bristol | VA | 51520 | Part of a bi-state metropolitan area with Tennessee |
| Portsmouth | VA | 51740 | Naval Medical Center Portsmouth |

**Table S3:**
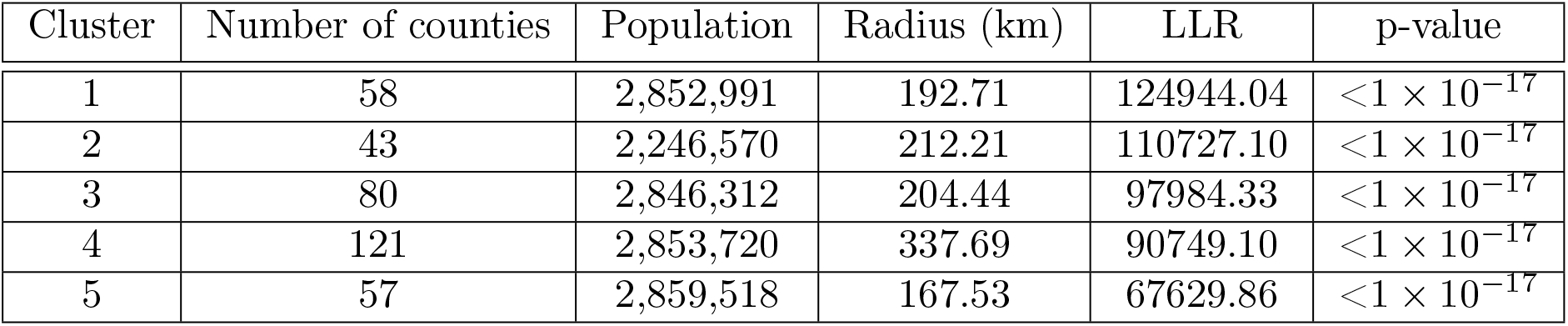
Additional information on the five most likely undervaccinated clusters. We provide data on the size and general reach of the clusters. We also give the log-likelihood ratio (LLR) and p-values.

| Cluster | Number of counties | Population | Radius (km) | LLR | p-value |
| --- | --- | --- | --- | --- | --- |
| 1 | 58 | 2,852,991 | 192.71 | 124944.04 | $<1 \times 10^{-17}$ |
| 2 | 43 | 2,246,570 | 212.21 | 110727.10 | $<1 \times 10^{-17}$ |
| 3 | 80 | 2,846,312 | 204.44 | 97984.33 | $<1 \times 10^{-17}$ |
| 4 | 121 | 2,853,720 | 337.69 | 90749.10 | $<1 \times 10^{-17}$ |
| 5 | 57 | 2,859,518 | 167.53 | 67629.86 | $<1 \times 10^{-17}$ |

## Notes

### Competing Interest Statement

The authors have declared no competing interest.

### Funding Statement

Research reported in this publication was supported by the National Institute of General Medical Sciences of the National Institutes of Health under award number R01GM123007. The content is solely the responsibility of the authors and does not necessarily represent the official views of the National Institutes of Health.

### Author Declarations

This research was deemed exempt by the Georgetown University Institution Review Board.

### Summary of Updates

Updated modeling process description for clarity, along with addition of marginal posterior plots; added additional references surrounding measles vaccine refusal; updated Figure S9 (formerly Figure S8)

